## Supplementary Figures for "Patterns of smartphone typing performance by time awake in US training physicians: implications for unobtrusive ambulatory mental fatigue assessment"

Yu Fang, et al.

**Figure S1.** SensorKit Keyboard Usage data collection.

**Figure S2.** The distributions of (A) time awake, (B) typing speed, and (C) rate of deletion in the sample.

**Figure S3.** Average rate of deletion of two example subjects by hours awake.

**Figure S4.** Average typing speed (A) and average rate of deletions (B) by number of hours awake, US English input mode only.

**Figure S5.** Average typing speed versus hours awake by the type of wearable device recording sleep.
**Figure S6.** Average rate of deletion versus hours awake, by the type of wearable device recording sleep.

**Figure S1.** SensorKit Keyboard Usage data collection.

|  | **DATA COLLECTED** | **DATA NOT COLLECTED** |
| --- | --- | --- |
| **SensorKit Keyboard Metrics** | - Length of words - Typing speed - Typing accuracy and the kind of typing errors - Keyboard orientation and screen size - Number of words and emoji typed | - Any of the words typed - Any information about keyboard extensions or stickers |

**Figure S2.** The distributions of (A) time awake, (B) typing speed, and (C) rate of deletion in the sample.

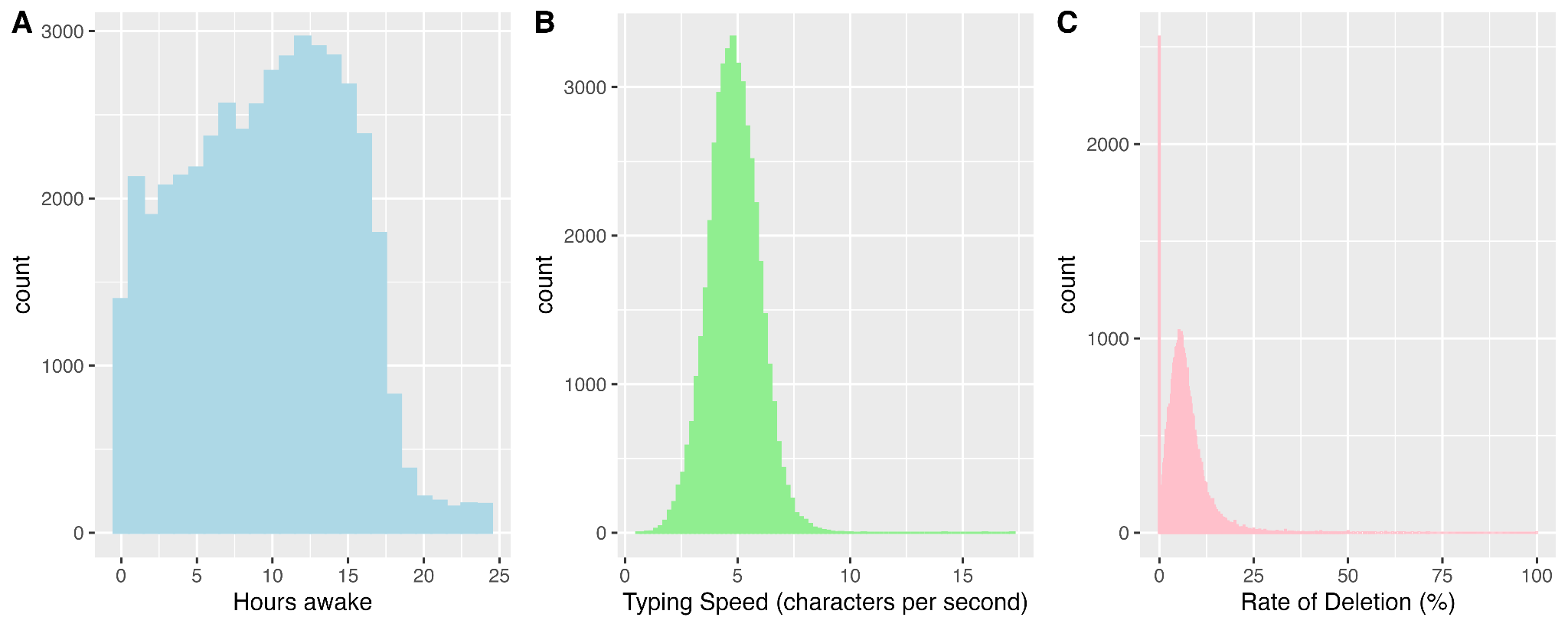

**Figure S3. Average rate of deletion of two example subjects by hours awake.** Rates of deletion were converted to individual z-scores. The Blue solid line represents the individual average (z-score = 0), while the red dashed line represents +0.1 standard deviation above the individual average.

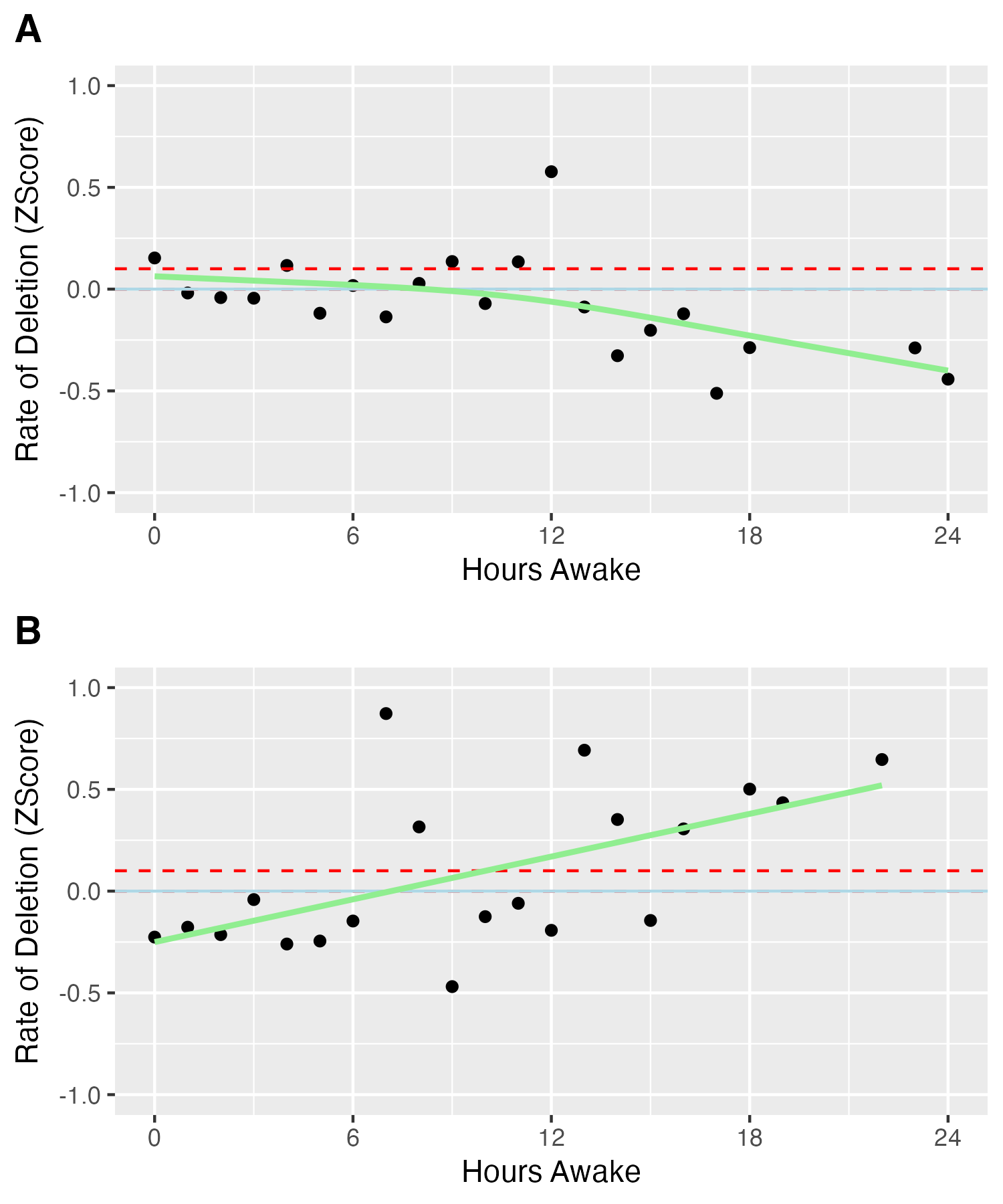

**Figure S4. Average typing speed (A) and average rate of deletions (B) by number of hours awake, US English input mode only.** Typing speed and rate of deletions were converted to individual z-scores. The error bars represent 95%CI. The GAM-smoothed prediction lines from model (4) were plotted (green). The Blue solid line represents the individual average (z-score = 0), while the red dashed line represents -0.1 standard deviation in (A) and +0.1 standard deviation in (B).

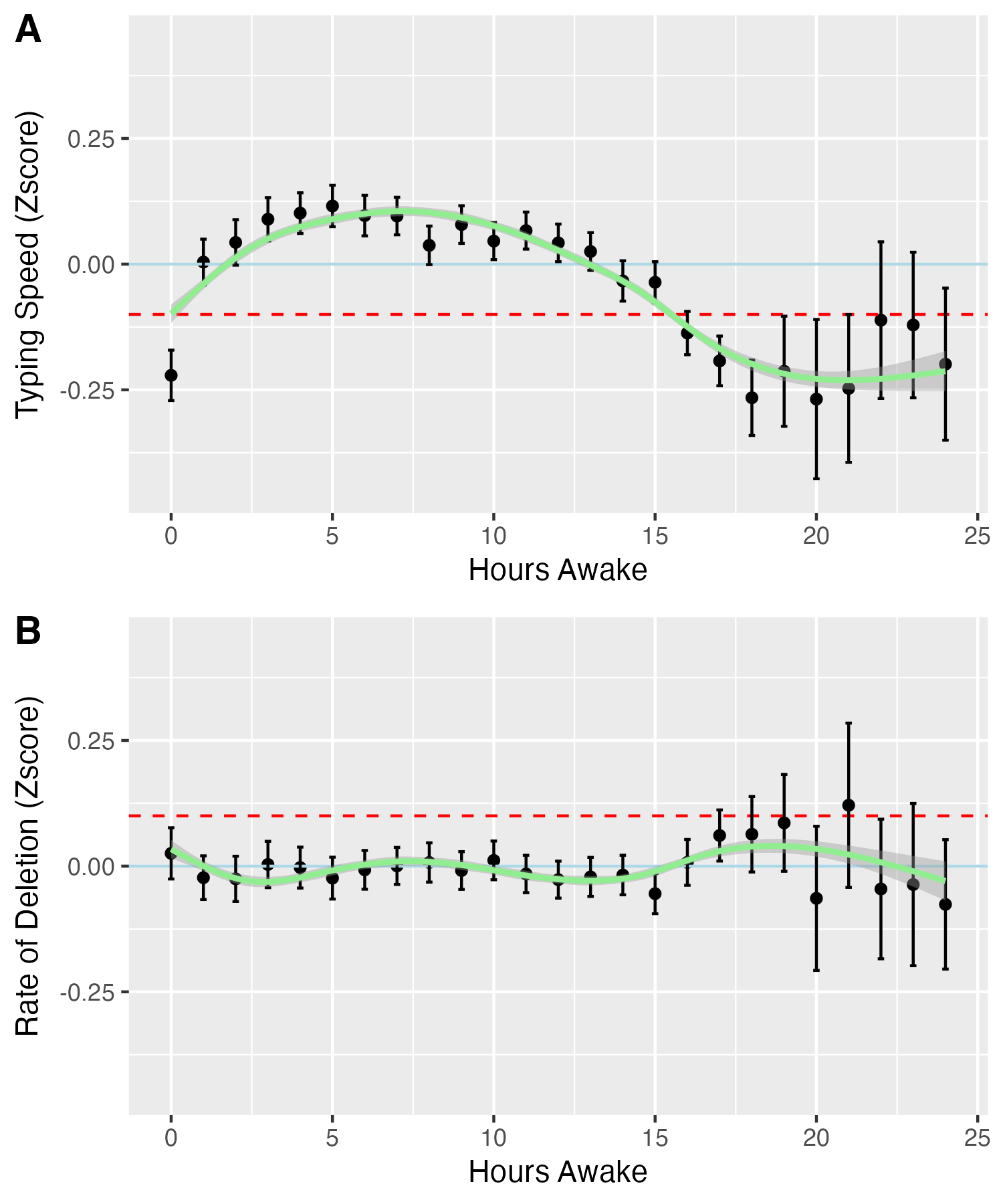

**Figure S5. Average typing speed versus hours awake by the type of wearable device recording sleep.** Typing speeds were converted to individual z-scores. The error bars represent 95%CI. The GAM-smoothed prediction lines from model (4) were plotted (green). The Blue solid line represents the individual average (z-score = 0), while the red dashed line represents -0.1 standard deviation.

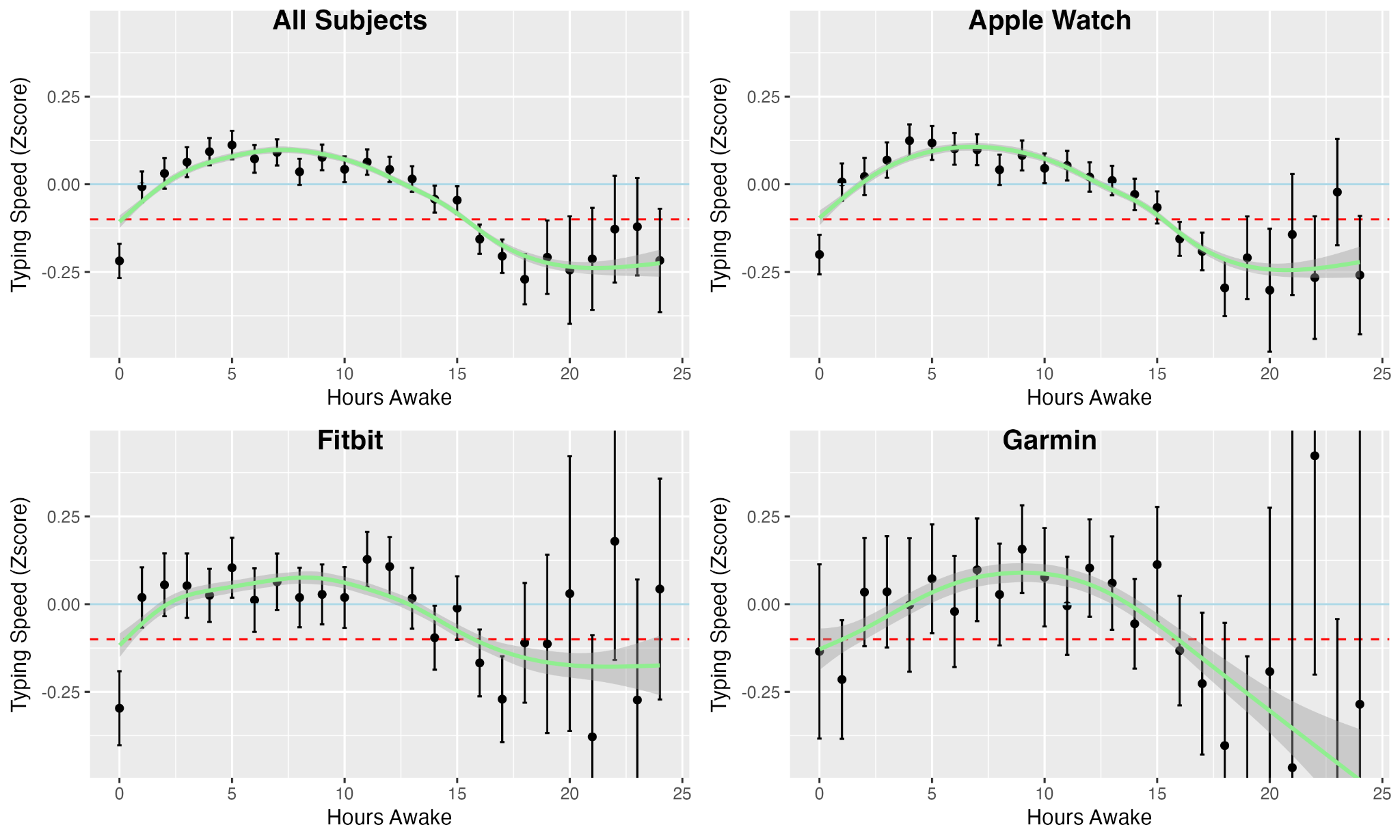

**Figure S6. Average rate of deletion versus hours awake, by the type of wearable device recording sleep.** Rates of deletions were converted to individual z-scores. The error bars represent 95%CI. The GAM-smoothed prediction lines from model (4) were plotted (green). The Blue solid line represents the individual average (z-score = 0), while the red dashed line represents 0.1 standard deviation.

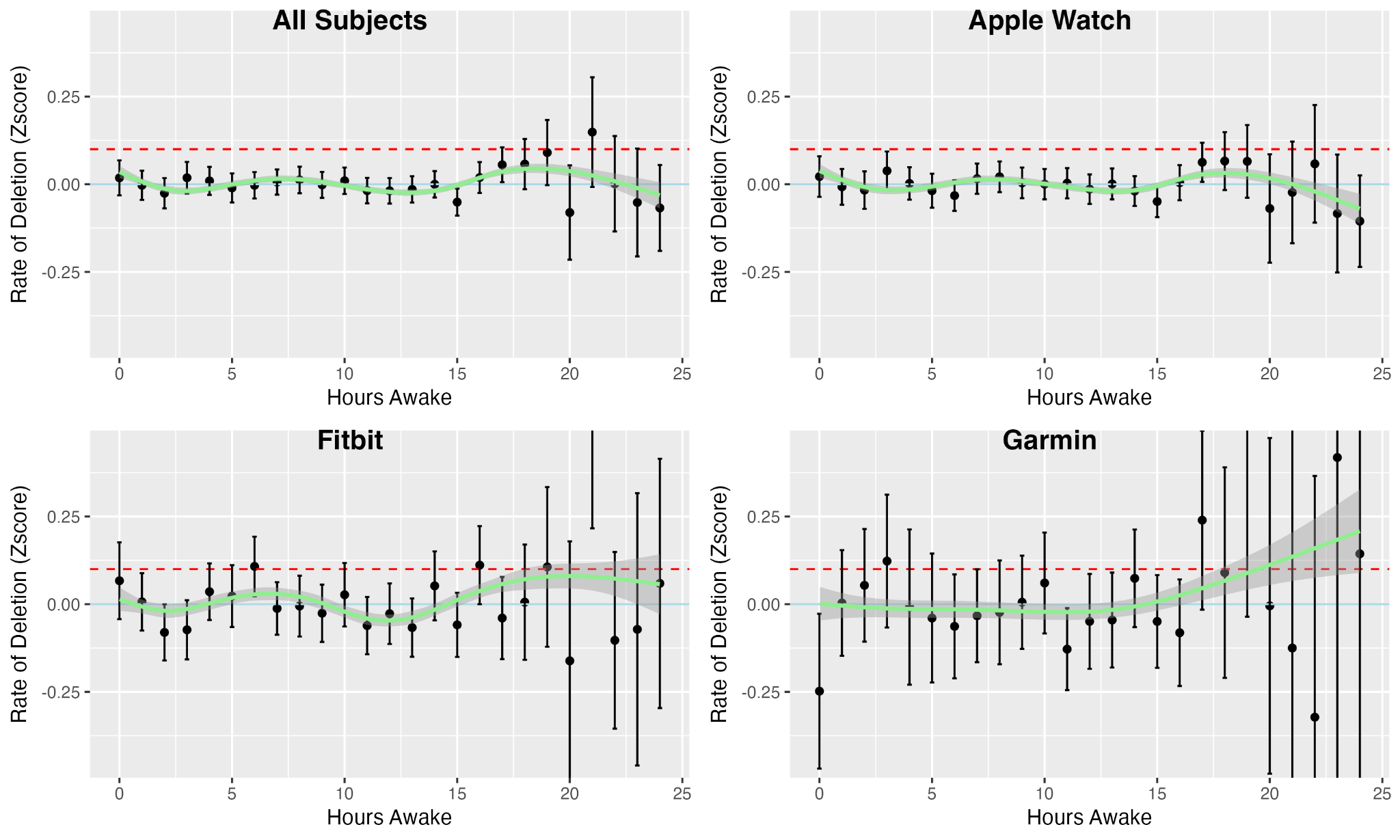
